# SARS-CoV-2 seroprevalence in the city of Hyderabad, India in early 2021

**DOI:** 10.1101/2021.07.18.21260555

**Authors:** Avula Laxmaiah, Nalam Madhusudhan Rao, N. Arlappa, Jagjeevan Babu, P. Uday Kumar, Priya Singh, Deepak Sharma, V. Mahesh Anumalla, T. Santhosh Kumar, R. Sabarinathan, M. Santhos Kumar, R. Ananthan, P.P.S. Blessy, D. Chandra Kumar, P. Devaraj, S. Devendra, M. Mahesh Kumar, Indrapal I. Meshram, B. Naveen Kumar, Paras Sharma, P. Raghavendra, P. Raghu, K. Rajender Rao, P. Ravindranadh, B. Santosh Kumar, Sarika, J. Srinivasa Rao, M.V. Surekha, F. Sylvia, Deepak Kumar, G. Subba Rao, Karthik Bharadwaj Tallapaka, Divya Tej Sowpati, Surabhi Srivastava, Manoj Murekhar, Rajkumar Hemalatha, Rakesh K Mishra

## Abstract

**Background:** COVID-19 emerged as a global pandemic in 2020, rapidly spreading to most parts of the world. The proportion of infected individuals in a population can be reliably estimated via sero-surveillance, making it a valuable tool for planning control measures. We conducted a serosurvey study to investigate SARS-CoV-2 seroprevalence in the urban population of Hyderabad at the end of the first wave of infections.

**Methods:** The cross-sectional survey conducted in January 2021 included males and females aged 10 years and above, selected by multi-stage random sampling. 9363 samples were collected from 30 wards distributed over 6 zones of Hyderabad and tested for antibodies against SARS-CoV-2 nucleocapsid antigen.

**Results:** Overall seropositivity was 54.2%, ranging from 50-60% in most wards. Highest exposure appeared to be among 30-39y and 50-59y olds, with women showing greater seropositivity. Seropositivity increased with family size, with only marginal differences among people with varying levels of education. Seroprevalence was significantly lower among smokers. Only 11% of the survey subjects reported any COVID-19 symptoms, while 17% had appeared for Covid testing.

**Conclusion:** Over half the city’s population was infected within a year of onset of the pandemic. However, ∼46% people were still susceptible, contributing to subsequent waves of infection.

**Highlights:**

⍰ National level serosurveys under-estimate localised prevalence in dense urban areas
⍰ SARS-CoV-2 seroprevalence in Hyderabad city was 54.2% after the first wave
⍰ A large proportion of the population remains at risk over a year into the pandemic

## Introduction

Coronavirus disease (COVID-19) was declared a pandemic over a year ago, when the infection due to severe acute respiratory syndrome coronavirus 2 (SARS-CoV-2) had spread worldwide [1]. During the first year of the COVID-19 pandemic, nearly 90 million cases were reported globally, with ∼2 million deaths [2]. Serological studies estimating SARS-CoV-2 exposure in human populations suggest that the true number of SARS-CoV-2 infections may be much higher than the officially reported cases [3]. This can be attributed to various factors, including the occurrence of asymptomatic infections, variable seeking of health care for clinically mild cases, varied testing strategies in different countries, false-negative virological tests, and incomplete case reporting.

Case reporting depends on several factors, including testing capacity, type of tests used, testing strategies, and health-seeking behaviour of the population. Many SARS-CoV-2 infections are mild or asymptomatic in nature and are less likely to be detected by the surveillance system. Therefore, population-based serosurveys are considered a valuable tool in estimating the proportion of the population infected with SARS-CoV-2. Another important use of serosurveys is to understand the demographic profiles of those at a higher risk of infection in different population groups. Large-scale population-based serosurveys are resource-intensive, and allocating scarce public health resources for this purpose could be challenging for many developing nations. Therefore, well designed population-based studies, with probability sampling and laboratory assays allowing high sensitivity and specificity followed by appropriate data analysis, play a crucial role in estimating the prevalence of the infected and susceptible populations [4].

Serosurveys conducted in Mumbai [5], Chennai [6] and Karnataka [7] have shown that in urban areas, the seroprevalence is much higher than that estimated at the national level. The Telangana media bulletin [8] revealed that the positivity rate in Telangana was 6.2%, case fatality rate was 0.58% and recovery rate was 85.9% in October 2020. Almost one-third of the cases were from Greater Hyderabad Municipal Corporation (GHMC) areas. The recent ICMR national sero-surveillance study (December 2020) reported a prevalence of 21.5% as against 12.2% in August and 0.33% during May 2020 [9,10]. Though this survey included 3 districts of Telangana, the estimates are indicative of national level prevalence. At state level, the sample size was too small to be representative and no sample was drawn from the GHMC area, which has a population of 10.3 million, Hyderabad being the fourth most populous city in India.

Considering the urgent need to estimate SARS-CoV-2 exposure in Hyderabad over the first year of the pandemic, we conducted a community-based seroprevalence study to assess the transmission of SARS-CoV-2 infection in the GHMC area. Our analysis estimates SARS-CoV-2 seroprevalence in the general population of Hyderabad, the socio-demographic risk factors for infection, and the trend of infectivity among various age groups, locations and socio-economic backgrounds in the city of Hyderabad.

## Methodology

### Study Design and sample size

The cross-sectional survey covered individuals aged 10 years and above. Assuming 10% seropositivity [9], a relative precision of 20%, confidence interval of 95%, design effect of 2.5, and non-response rate of 20%, we estimated a sample size of 2593 (rounded to 3,000) individuals in each age category to be effective. To carry out segregated analysis across age groups (≥10 years to 18 years, 18 years to 60 years and above 60 years), the survey was planned for a sample size of at least 9000 individuals from different locations (wards) in the city. The wards covered 6 different zones of Hyderabad, with the zones subdivided further into 19 circles (covering 30 Wards).

### Sampling procedure

About 30 wards were selected using a simple random sampling technique from the list of 150 wards in the GHMC area. Each selected ward was tentatively divided into 4 segments and from each segment nearly 25 households were covered by selecting the first household randomly and covering the next 24 households contiguously. Thus, a total of around 100 households were sampled from every ward, including all consenting and available males and females aged ≥10 years from each household. Subjects were included in the survey irrespective of their current COVID status. Non-consenting subjects, debilitated, bed-ridden or severely sick subjects were excluded from the study.

### Data collection and ethics approvals

Data were collected by 15 field teams and 3 lab teams. Each field team consisted of a medical officer/scientist, a technician and a phlebotomist. Each lab team consisted of a scientist (microbiologist) and 3 lab technicians. For supervision and monitoring, there were 3 survey coordinators, one lab coordinator and one overall study coordinator. The data collection was completed in the month of January in two phases, i) 1^st^ phase: 8^th^-12^th^ January 2021 and ii) 2^nd^ Phase: 21-24^th^ January 2021. Data were collected through ODK based Computer Assisted Personal Interview with a structured questionnaire which had mostly closed ended questions. The study team visited the randomly selected households and briefed them about the survey objectives and the process involved. The state health department and other authorities actively participated to cooperate with the survey teams. An informed written individual consent was taken from all participants. Subjects 10-18 years of age were asked for consent which was counter signed by parents/legal guardians. A participant information sheet was also provided to each participant elucidating the study details. Interviews were conducted at the households as per the convenience of the participants in order to ensure privacy.

After obtaining written individual informed consent, information on socio-economic and demographic details, exposure history to laboratory confirmed COVID-19 cases, symptoms suggestive of COVID-19 since the beginning of the pandemic, and clinical history were recorded. When there was unavailability of eligible individuals in a household, the data collection team moved onto the next household for enrolling the required number of subjects. Trained phlebotomists in each of the 15 survey teams collected 3-4 ml of venous blood from each participant. Serum was separated after centrifugation at ICMR-NIN. Estimation of SARS-CoV-2 specific antibodies was performed at the CSIR-CCMB, Hyderabad. Data were stored securely under the investigator’s responsibility, with a focus on ensuring the participant’s confidentiality. Samples were anonymized and except the principal investigator, the identifying details were not shared with anyone. However, the IgG antibody test results were shared with each individual for their information. Final reports and aggregated data were prepared without any identifying information.

### Antibody titre assays and measurement

The samples were tested for total SARS-CoV-2 antibodies via electrochemiluminescence immunoassay using Elecsys Anti-SARS-Cov2 kit (Roche Cobas E411) based on a recombinant protein representing the nucleocapsid (N) antigen for antibody determination, as per manufacturer’s protocol. Samples that had >1 COI (cutoff index; signal sample/cutoff) were considered positive for presence of antibodies.

### Statistical and data analysis

In-house scripts were used for filtering and categorization of data. Samples with unknown data fields were removed from analysis. Seroprevalence was calculated based on the number of samples with antibody titre >1 COI and analyzed according to the demographic measures surveyed. For analysis of family transmission, households with only one member or those with no seropositive members were excluded. Data was analysed and visualised using SPSS v.22 and ggplot2. Correlation was calculated using the Pearson coefficient. Significance of association between various groups and seropositivity was assessed using Chi-square test.

## Results

### Over half the surveyed population showed prior exposure to SARS-CoV-2 infection

A total of 9517 blood samples were collected from 4456 households, residing in 30 wards distributed over 6 zones across the city of Hyderabad, India (see Methods). 154 samples were rejected or had incomplete metadata and seroprevalence was assessed from the remaining 9363 samples. Of these, 5076 were positive for the SARS-CoV-2 antibodies thus giving an overall positivity of 54.2% (95% CI: 53.2-55.2). Most of the wards surveyed had a uniform distribution of seropositivity ranging from 50-60% (Figure 1). However, a few wards showed evidence of higher exposure to the coronavirus (maximum ward seroprevalence of ∼72%) while 8 wards had seroprevalence <50%, indicating a more susceptible population in these areas. Out of the 6 zones making up the city, Secunderabad had the highest seroprevalence of 61.6%, while L.B. Nagar showed the lowest seroprevalence of 43.3%.

**Figure 1.**
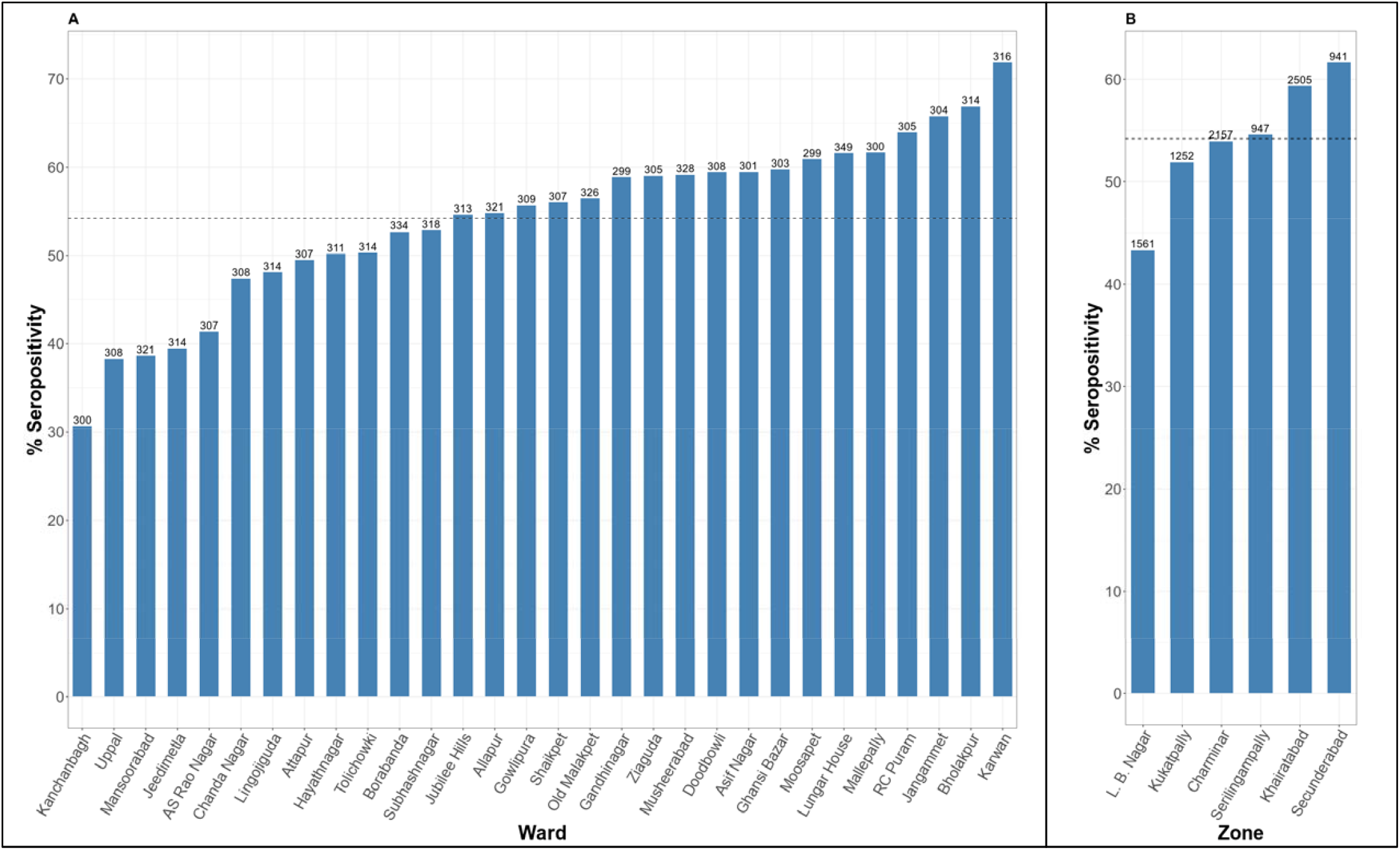
Estimated seroprevalence across different GHMC wards in Hyderabad. Seropositivity % (Y-axis) plotted across A) 30 wards and B) 6 zones in Hyderabad (9363 individuals surveyed). We find an average positivity of 54.2% (dotted line, 95% CI: 53.2-55.2). Most of the wards surveyed had a uniform distribution ranging from 50-60% seropositivity. Values on the top of the bars indicate number of individuals surveyed in the group.

### Prevalence among various socioeconomic and demographic groups

The socioeconomic status and demographics of the participants are summarised in Table 1. The study participants were ≥10 years of age (mean age = 36.6 years, SD = 16.4) and were grouped into 7 bins as shown in Figure 2. The lowest seroprevalence was seen among people in the age group >70 years (47.6%, p <0.05, CI 95%), possibly reflecting a poorer geriatric immune response or lower mobility and/or a greater degree of precautions taken by older individuals during the pandemic. The highest exposure was in the age group of 30-39 and 50-59 year olds (56.7%). The number of individuals in their 20s and 30s sampled was the highest (43% of total respondents), while there was a lower representation of older individuals (349 samples, grouped together in the > 70 years age group).

**Table 1.** Demographic Characteristics of study subjects.

| **Variable** |  | **%** |
| --- | --- | --- |
| **Gender** | Male | 44.94 |
|  | Female | 54.92 |
|  | Other Gender | 0.14 |
| **Age Group (Years)** | 10 to 19 | 16.47 |
|  | 20-29 | 21.53 |
|  | 30-39 | 21.49 |
|  | 40-49 | 16.83 |
|  | 50-59 | 12.09 |
|  | 60-69 | 7.86 |
|  | 70 & Above | 3.73 |
| **Education Levels** | Illiterate | 15.73 |
|  | Read & Write | 9.04 |
|  | Primary | 14.64 |
|  | Secondary | 24.76 |
|  | Intermediate | 13.49 |
|  | Graduation & Above | 22.34 |
| **Family Size** | <=4 | 65.12 |
|  | 5 to 6 | 29.30 |
|  | >=8 | 5.58 |
| **Number of Rooms in their residence** | <=2 | 51.21 |
|  | 3 to 4 | 41.91 |
|  | 5 to 6 | 5.94 |
|  | >=7 | 0.94 |

**Figure 2.**
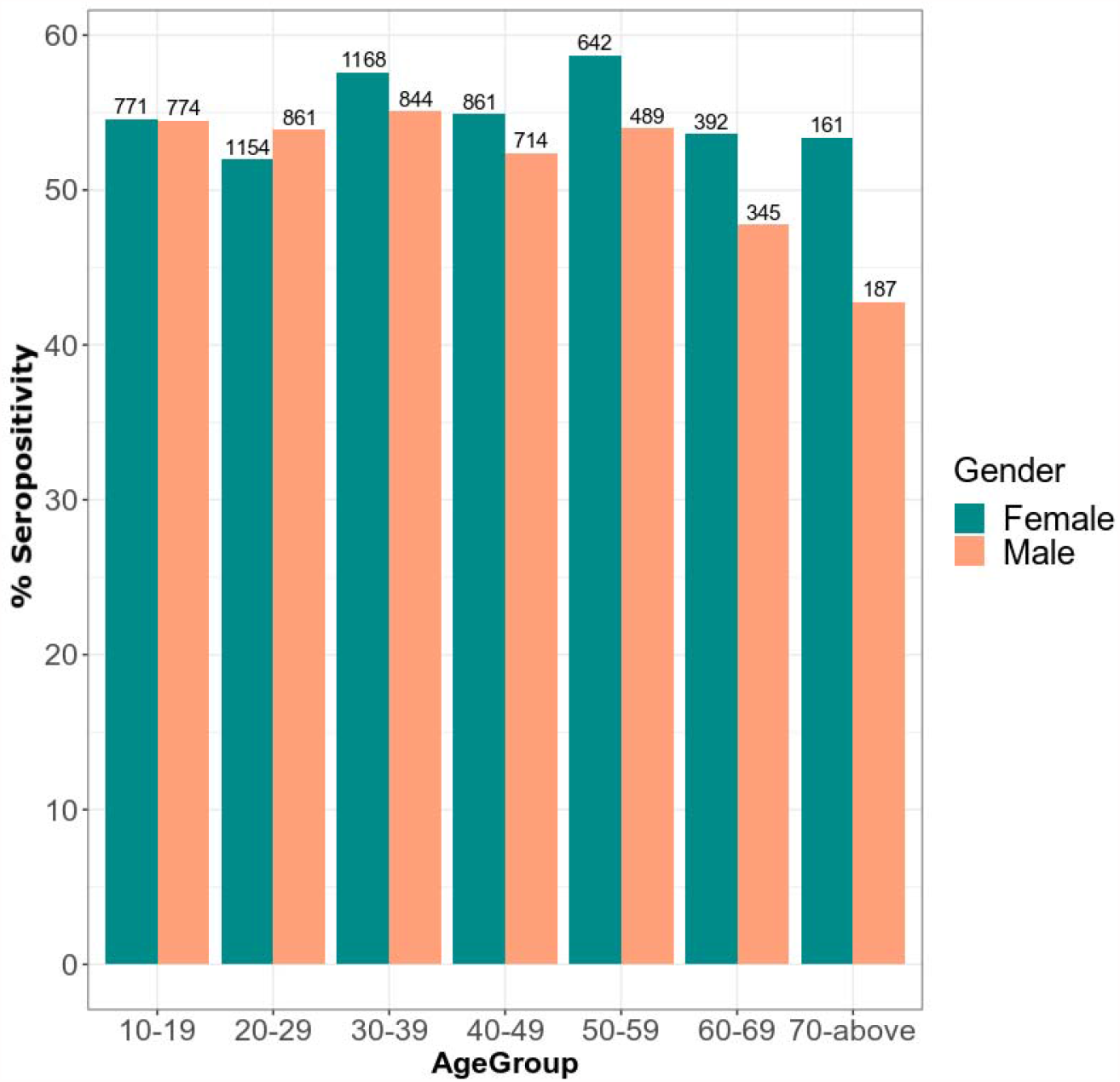
Seroprevalence (%) of SARS-CoV-2 by age and gender groups. Seropositivity % (Y-axis) plotted across demographic groups. The study participants (≥10 years of age; mean age = 36.6 years, SD = 16.4) were binned into 7 age-groups of a decade each, by gender (55% females). Values on the top of the bars indicate number of participants in the group.

Approximately 55% of the samples were from females. Enrolled individuals consisted of 5143 females and 4209 males and the weighted seroprevalence was marginally higher in females (55.2%, 95% CI, 53.8-56.6%) compared to males (53.0%, 95% CI, 51.5-54.5%). This difference was statistically significant (p=0.036) and the same trend was seen across all the age groups, except in individuals between 10 to 29 years (Figure 2 and Table 2).

**Table 2.** Prevalence of SARS-CoV-2 antibodies among people of GHMC, Telangana by sample characteristics.

| **Variable** |  | **Positive cases** | **% Sero-positivity** | **Total Number** | **P value** |
| --- | --- | --- | --- | --- | --- |
| **Gender** | Male | 2231 | 53.0 | 4208 | 0.036 |
|  | Female | 2838 | 55.2 | 5142 |  |
|  | Other Gender | 7 | 53.8 | 13 |  |
|  | Total | 5076 | 54.2 | 9363 | 0.004 |
| **Age Group (Years)** | 10-19 | 842 | 54.6 | 1542 |  |
|  | 20-29 | 1063 | 52.7 | 2017 |  |
|  | 30-39 | 1141 | 56.7 | 2012 |  |
|  | 40-49 | 847 | 53.7 | 1575 |  |
|  | 50-59 | 642 | 56.7 | 1132 |  |
|  | 60-69 | 375 | 51.0 | 736 |  |
|  | 70 & Above | 166 | 47.6 | 349 |  |
|  | Total | 5076 | 54.2 | 9363 |  |
| **Education Levels** | Illiterate | 819 | 55.6 | 1473 | 0.0001 |
|  | Read & Write | 473 | 55.8 | 847 |  |
|  | Primary | 723 | 52.7 | 1371 |  |
|  | Secondary | 1334 | 57.5 | 2318 |  |
|  | Intermediate | 698 | 55.3 | 1263 |  |
|  | Graduation & Above | 1029 | 49.2 | 2091 |  |
|  | Total | 5076 | 54.2 | 9363 | 0.005 |
| **Family Size** | <=4 | 3237 | 53.1 | 6098 |  |
|  | 5-7 | 1532 | 55.9 | 2743 |  |
|  | >=8 | 307 | 58.8 | 522 |  |
|  | Total | 5076 | 54.2 | 9363 |  |
| **Number of Rooms in their residence** | <=2 | 2635 | 54.9 | 4796 | 0.216(NS) |
|  | 3-4 | 2106 | 53.7 | 3924 |  |
|  | 5-6 | 295 | 53.2 | 555 |  |
|  | >=7 | 40 | 45.5 | 88 |  |
|  | Total | 5076 | 54.2 | 9363 |  |
| **History of Contact with COVID -19 subjects** | Yes(In HH) | 189 | 78.4 | 241 | 0.0001 |
|  | Yes(Outside HH) | 228 | 67.3 | 339 |  |
|  | No | 3517 | 52.6 | 6688 |  |
|  | Don't Know | 1142 | 54.5 | 2095 |  |
|  | Total | 5076 | 54.2 | 9363 |  |
| **COVID-19 Test Status** | Test Positive | 260 | 87.2 | 298 | 0.0001 |
|  | Test Negative | 713 | 55.2 | 1291 |  |
|  | Total | 973 | 61.2 | 1589 |  |

Nearly 84% of the participants were literate with the most common level of education being secondary school (24.75%) or graduation and above (22.3%). SARS-CoV-2 exposure levels were similar across the various educational strata, ranging between 52.7% and 57.5%, but only among the graduates, it remained low at 49.2% (p<0.001). The exposure levels also showed an increasing trend with the family size, ranging from 53.1% when the family size was up to 4 people and 58.8% when it was 8 or more (p<0.05) (Table 2).

### Degree of transmission in household

A majority (71.4%) of the individuals reported no known contact with COVID positive persons, yet 52.6% of them were seropositive (with possibly unknown source of infection), similar to the overall population prevalence (Table 2). 3.61% individuals reported contact with a known COVID positive person outside their own household, and of these 67.3% were found to be seropositive. Only 2.57% of the total participants reported contact within their household and the seropositivity was found to be the highest (78.4%) within this group, suggestive of effective family transmission.

In order to estimate the degree of household transmission, we analyzed the family members across all the households surveyed. Families consisted of 1-9 members, living in single or multi-room homes ranging from 1 to >5 rooms per household. Majority of the households surveyed constituted small families with 4 or fewer members (65.1%). Nearly half of the surveyed households dwelt in houses with only 2 rooms (51.2%). 1473 households had no seropositive family members and were not considered for the family transmission analysis. Among families where at least one member was seropositive, no specific trends could be observed with increasing room number or space for isolation in the context of avoiding infection spread.

### Correlation with confirmed COVID-19 or other diseases

Though more than half the population was positive in the antibody assay, indicating a prior exposure to SARS-CoV-2, very few individuals appeared for COVID testing (17% of our study group) (Figure 3a). Importantly, 87.2% of the COVID test positive individuals (either Rapid Antigen Test or RT-PCR test positives) still had detectable antibodies to the virus, suggesting retention of the antibody response at the time of this study (Figure 3b), compared to only ∼53% of those who were not tested, or 55% of those who were negative for the COVID test. However, since the precise dates of the COVID testing were not available, the period of the retention of antibody response cannot be estimated from this study.

**Figure 3.**
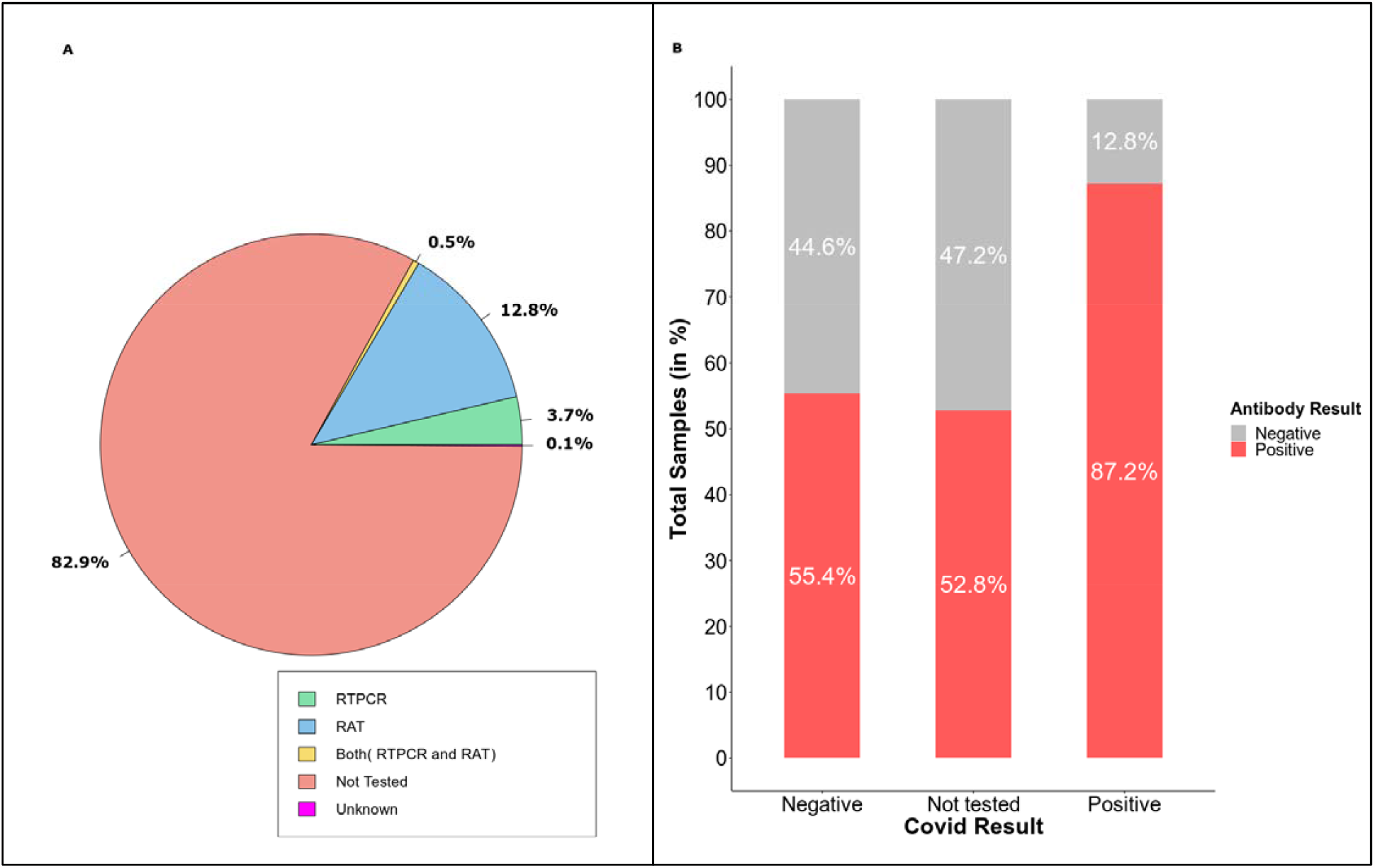
Distribution of survey subjects by Covid testing status. A) Pie-chart representing percentage of participants who appeared for a Covid test (RAT and/or RT-PCR) and B) Percentage of individuals (Y-axis) seropositive (red) or not (grey) categorized by their Covid test result (X-axis).

We also looked for their symptoms status, and found that only about 11% of the total individuals surveyed (1009 out of 9363) reported any of the symptoms known to be associated with COVID-19 (Figure 4). These results suggest that most of the seropositive people were unaware of having contracted the infection and a majority of them remained asymptomatic. As expected, the seropositivity was higher in the symptomatic group (61.7%) compared to the asymptomatic group.

**Figure 4.**
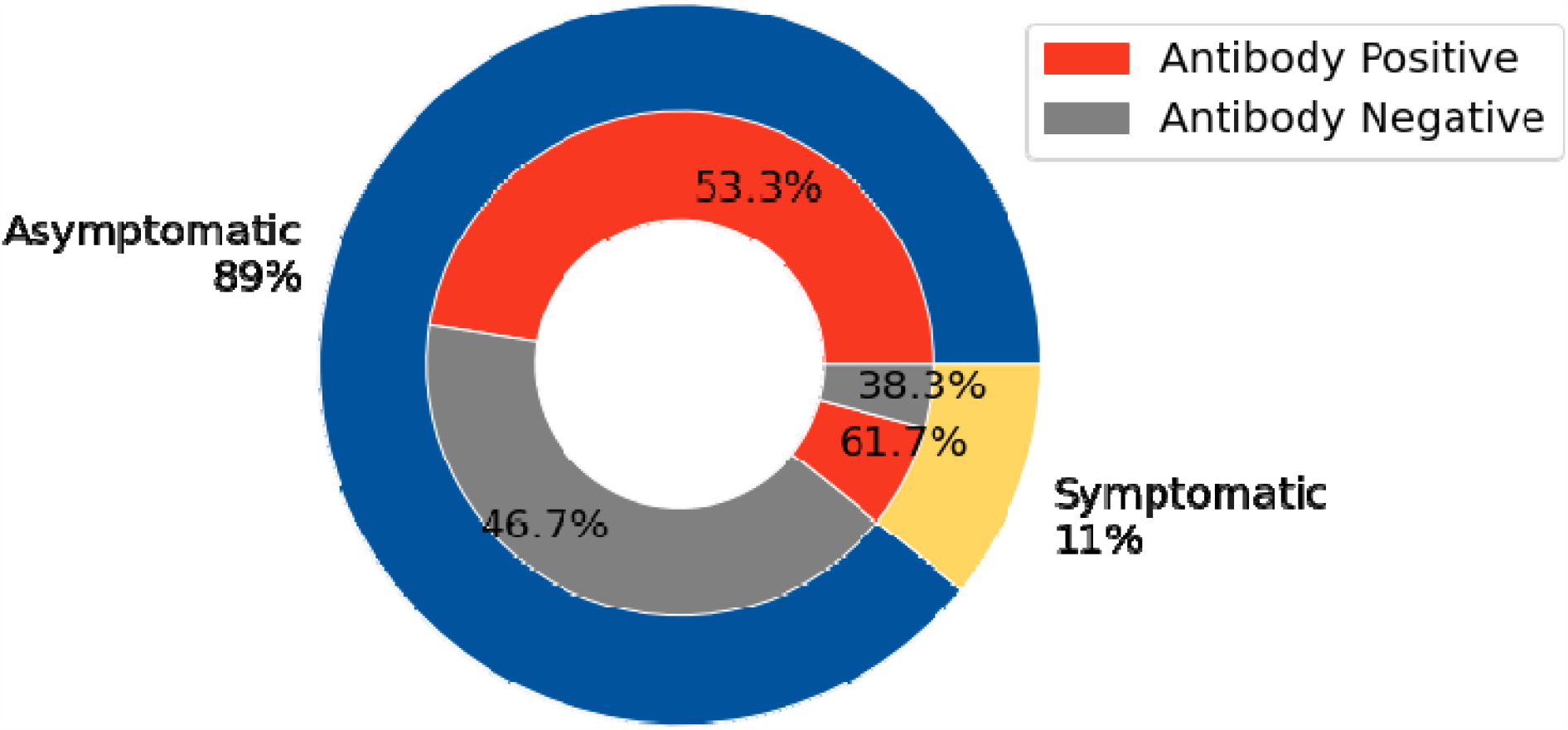
Seroprevalence (%) of SARS-CoV-2 by symptom status. Donut chart showing higher % seropositivity (red, inner pie) in context of individuals who showed COVID-19 related symptoms compared to those who did not (symptomatic and asymptomatic, respectively, outer pie).

Among the eight symptoms covered in our survey (Table 3), cough and fever were reported by nearly 550 (5.87%) individuals while diarrhea, excessive tiredness, sore throat, and loss of smell and taste were reported by very few individuals (<340). Loss of smell and taste, however, showed the strongest association with seropositivity among the few individuals who reported these symptoms (>86%). Among those reporting the relatively more common symptoms of cough and fever, seroprevalence was also found to be higher than the population average (ranging from 61-72.5%). Most of the symptomatic individuals suffered from only one or two symptoms (487/1009, 48.3% and 328/1009, 32.5%, respectively) while a few reported a combination of 3 or more symptoms (194/1009, 19.2%).

**Table 3.**
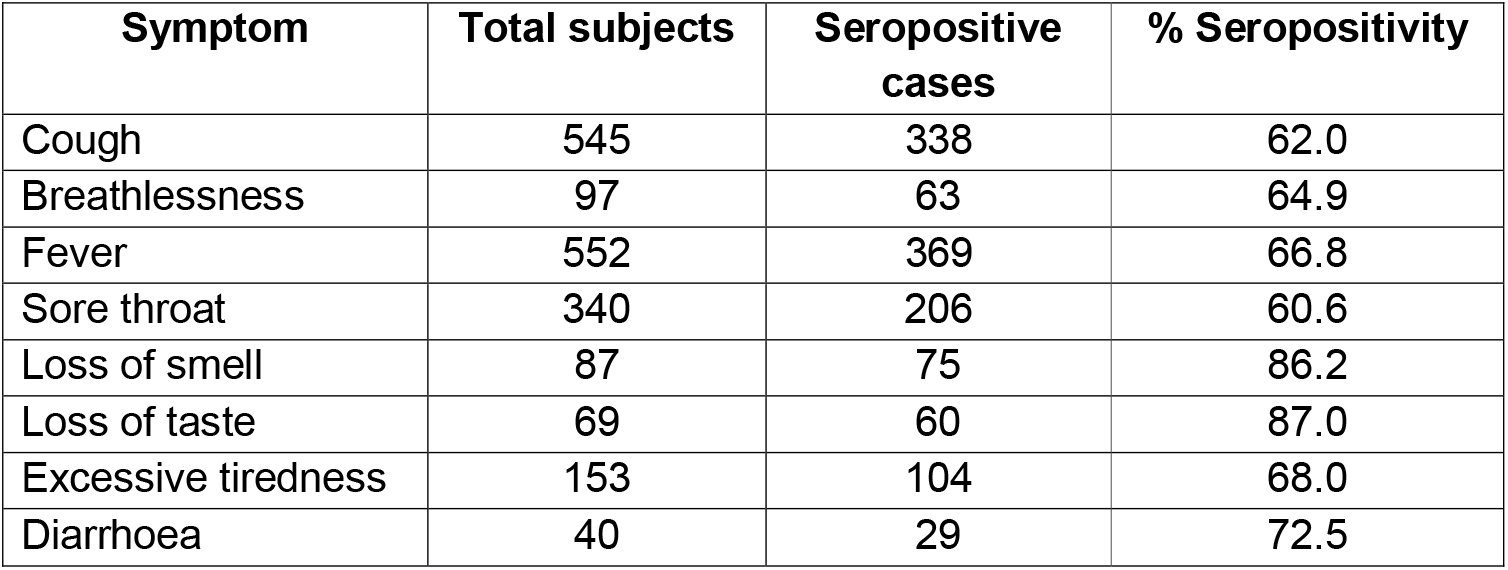
Symptoms presented by the study subjects.

| **Symptom** | **Total subjects** | **Seropositive cases** | **% Seropositivity** |
| --- | --- | --- | --- |
| Cough | 545 | 338 | 62.0 |
| Breathlessness | 97 | 63 | 64.9 |
| Fever | 552 | 369 | 66.8 |
| Sore throat | 340 | 206 | 60.6 |
| Loss of smell | 87 | 75 | 86.2 |
| Loss of taste | 69 | 60 | 87.0 |
| Excessive tiredness | 153 | 104 | 68.0 |
| Diarrhoea | 40 | 29 | 72.5 |

Very few participants reported being afflicted with comorbidities or other systemic diseases associated with increased severity of COVID-19; 78% of the subjects had none of the 8 comorbidities tested (Table 4). Further, even among the individuals with more prevalent comorbidities such as diabetes and hypertension (1623 individuals), there was no change in seropositivity, which remained at 54.1%. We found lower seropositivity (40%) among self-declared smokers compared to the non-smokers. Although the number of participants who smoked was small (275), these results were significant (p<0.05, 95% CI) and suggest possible protection against COVID-19. It remains to be established if there are any behavioural links that reduced the chance of infection in this study group or whether they have poorer or shorter duration of antibody response.

**Table 4.**
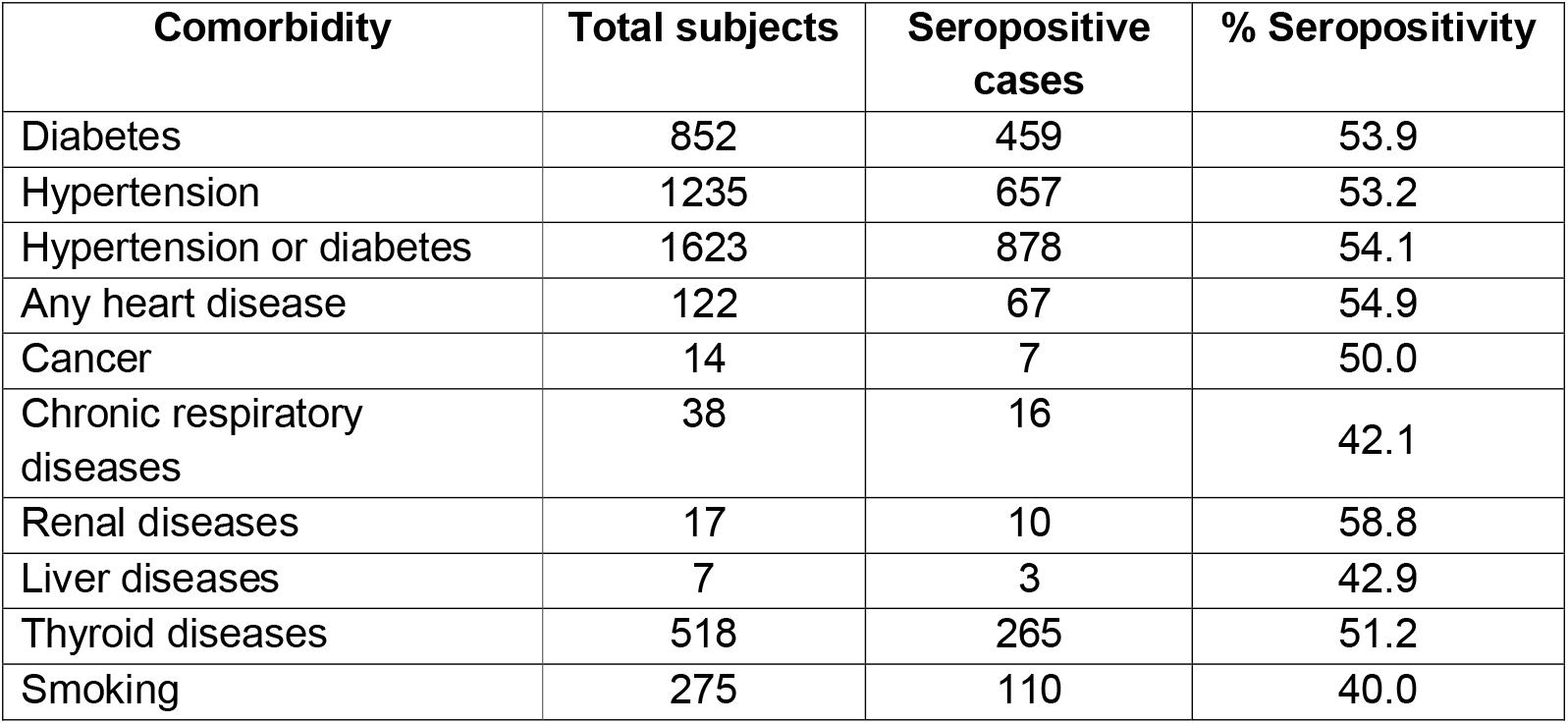
Comorbidities reported by the study subjects.

| **Comorbidity** | **Total subjects** | **Seropositive cases** | **% Seropositivity** |
| --- | --- | --- | --- |
| Diabetes | 852 | 459 | 53.9 |
| Hypertension | 1235 | 657 | 53.2 |
| Hypertension or diabetes | 1623 | 878 | 54.1 |
| Any heart disease | 122 | 67 | 54.9 |
| Cancer | 14 | 7 | 50.0 |
| Chronic respiratory diseases | 38 | 16 | 42.1 |
| Renal diseases | 17 | 10 | 58.8 |
| Liver diseases | 7 | 3 | 42.9 |
| Thyroid diseases | 518 | 265 | 51.2 |
| Smoking | 275 | 110 | 40.0 |

## Discussion

The Indian Council of Medical Research (ICMR) has been carrying out repeated cross-sectional surveys in 70 districts from 21 states for national level estimation and 3 rounds of surveys have already been completed and reported. Since these surveys are not sufficient to draw inferences at a micro level, the present survey was designed to estimate seroprevalence levels at the end of the first year of the pandemic in the GHMC area (Hyderabad, India), between the first and the second waves of infections.

Pan-India seroprevalence studies began in May 2020 in India (when the assumed prevalence was 1% or lower). The seroprevalence was, at its lowest, found to be 1% in the state of Kerala and at pan-India level in June [10], increasing to 19% in November [14]. We have documented much higher seroprevalence in Hyderabad (54.2%) than that seen in other states such as Tamil Nadu (31%) [15], Uttar Pradesh, Gujarat, West Bengal, Madhya Pradesh, Karnataka and Chhattisgarh (41%) [16] in the same time frame (November 2020 to January 2021). A few other studies have reported much lower seroprevalence of 17.6% in Ahmedabad [17] and 3.1% in Srinagar [18] in November-December 2020. The findings of our study are similar to those from Mumbai [5], Chennai [6] and in Karnataka [7] conducted during the months of July 2020, suggesting that urban populations have had much higher seroprevalence than the national average prevalent at that time. An important caveat to note is that antibody testing kits used in seroprevalence surveys aren’t very sensitive, nor are they all uniform and direct comparisons of results from different studies should be interpreted cautiously.

We found gender-specific differences in seropositivity levels as also documented by some of the above-mentioned serosurveys in India. Females appear to generate better protective antibody responses than do males following vaccination against influenza, yellow fever, dengue, and several other viruses [11]. Differential exposure and susceptibility, and behavioural and immunological divergence between the genders have been cited to account for higher seroprevalence found in other surveys in the country. A recent review looking at global seroprevalence rates concluded that in most other countries, however, males had a slightly higher seropositivity than females, or there was no difference found between the genders [12].

Though we found no correlation with any known comorbidities some difference in seroprevalence was seen with smoking, which causes the upregulation of ACE-2 receptor [13]. Most of the survey subjects appeared to be asymptomatic for the known COVID-19 symptoms prevalent in the first wave of infections, and were likely unaware of their infected status. It is unclear whether genetic and/or environmental and behavioural differences contribute to any of the observed differences among individuals. Larger studies in these target groups are needed for direct comparison and further conclusions.

A significant aspect of this study was the identification of pockets of low seroprevalence (as low as 31%) at the start of this year. These represent a reservoir of uninfected and susceptible individuals, that may have contributed to the large degree of cases seen in the second wave of infections across the country. With the ongoing vaccination drive expected to take many months to complete, and the emergence of novel variants of the virus that may have properties of immune escape or increased transmission, we cannot afford to let our guard down at this stage. Frequent serosurveys will be essential in monitoring the course of the pandemic in the months ahead.

## Conclusions and implications

This study shows that the overall SARS-CoV-2 seropositivity was about 54% among the population of GHMC, Hyderabad, Telangana, not including children below 10 years of age. It is highly desirable that, irrespective of the seropositivity levels seen in the beginning of this year, most eligible individuals get vaccinated, taking advantage of the available vaccines that provide robust protection. A high number of SARS-CoV-2 infections, as seen in the last couple of months, provide the replicating virus with a chance to acquire mutations with consequences for current pandemic mitigation strategies. In the worst case scenario, the benefits gained by high seroprevalence or the ongoing vaccination drive may be undone by emerging immune escape variants. It is, therefore, highly advisable to promote the continued use of non-pharmacological measures like wearing masks, hand hygiene and physical distancing while avoiding indoor and large-scale gatherings.

## Supporting information

STROBE cross-sectional checklist

## Data Availability

Anonymized participant data are available with the authors

## Funding

This work was supported by institutional funding of ICMR-NIN and CSIR-CCMB, with generous support from Bharath Biotech Pvt. Limited. Bharath Biotech Pvt. Limited was involved in procurement & supply of the test kits and had no role in design of the study, data handling, sample analysis or in the preparation of the manuscript. All authors had access to all the data in the study and had final responsibility for the decision to submit for publication.

Institutional funding of ICMR – National Institute of Nutrition, Hyderabad, and CSIR-Centre for Cellular and Molecular Biology, Hyderabad.

Bharath Biotech Pvt. Ltd., Hyderabad.

## Informed Consent

Written informed consent was obtained from all the survey participants. Samples were anonymized, and privacy of individual participants was maintained.

## Acknowledgement

Anitha Ch, Anjaiah N., Aruna Kumari T., Aruna Reddy V., Bhavani G., Chandrababu V., Chathyusha., Jhansi N., Juhika., Munikumar M., Madhu K., Sameera S., Nancharamma BV., Narasimhulu D., Nagender., Nasar vali SK., Panda H., Praveen B., Prasad SPV., Purnachandra., Ranjit Babu., Raji Reddy GV., Rajyalaxmi R., Rani D., Satyanarayan R., Saibabu C., Sathaiah P., Sarala D, Sriram P., Sreenu P., Sheela E., Sree Ramakrishna K., Swetha S., Swaroopa D., Suresh M., Srinivas D., Subhash., Venkatamma P., Vaishnavi., Venkataramana., Vijayalaxmi G., Tulsi Bai G, Tulja B, Usha T: helped in data, and blood sample collection.

## Author Contributions

**AL, NMR, SS:** Overall execution of the study and draft writing of manuscript

**NA, JJB, PUK, SS, DTS**: Supervision of data collection, quality control of data entry and critical review of the manuscript

**MAV, SKT, KBT**: Estimation of IgG antibodies from serum samples

**BDK**: Processing of blood and preparation of serum aliquots

**MVM, MSK, RSN**: Study methodology and development of ODK plot form online data collection

**NKB, PS, DS, SS, DTS:** Data processing, data analysis and visualization

**RA, BPPS, CKD, SD, MMK, IIM, BNK, PS, PRV, PR, KRR, PRN, BSK, GS, JSR, MVR, SF, BDK, GSR**: Supervision of teams, data collection and review of manuscript

**RH, RKM**: Overall coordination of the study and critical review of the manuscript

## Abbreviations

ACE2: Angiotensin-converting enzyme 2
CCMB: Centre for Cellular & Molecular Biology
CI: Confidence Interval
COVID-19: Coronavirus Disease
COI: Cutoff index
CSIR: Council of Scientific and Industrial Research
GHMC: Greater Hyderabad Municipal Corporation
ICMR: Indian Council of Medical Research
NIN: National Institute of Nutrition
ODK: Open Data Kit
RT-PCR: Reverse transcription polymerase chain reaction
SARS: Severe Acute Respiratory Syndrome
SD: Standard Deviation
SPSS: Statistical Package for the Social Sciences

## References

1. WHO. Coronavirus disease 2019 (COVID-19) Situation Report – 90 Available from: https://www.who.int/docs/default-source/coronaviruse/situation-reports/20200419-sitrep-90-covid-19.pdf?sfvrsn=551d47fd_2 (accessed on April 20, 2020)

2. WHO. 1.8 million global deaths as of 1^st^ week of Jan 2021, https://www.who.int/publications/m/item/weekly-epidemiological-update5-january-2021 (Accessed on April 20, 2020)

3. Chen X, Chen Z, Azman AS, Deng X, Sun R, Zhao Z, Zheng N, Chen X, Lu W, Zhuang T, Yang J, Viboud C, Ajelli M, Leung DT, Yu H. Serological evidence of human infection with SARS-CoV-2: a systematic review and meta-analysis. Lancet Glob Health. 2021 May;9(5):e598–e609.

4. Murhekar MV, Clapham H. COVID-19 serosurveys for public health decision making. Lancet Glob Health. 2021 May;9(5):e559–e560. doi: 10.1016/S2214-109X(21)00057-7.

5. Malani A, Shah D, Kang G, Lobo GN, Shastri J, Mohanan M, Jain R, Agrawal S, Juneja S, Imad S, Kolthur-Seetharam U. Seroprevalence of SARS-CoV-2 in slums versus non-slums in Mumbai, India. Lancet Glob Health. 2020 Nov 13:S2214-109X(20)30467-8.

6. Selvaraju S, Kumar M, Thangaraj J, Bhatnagar T, Saravanakumar V, Kumar C, et al. Population-Based Serosurvey for Severe Acute Respiratory Syndrome Coronavirus 2 Transmission, Chennai, India. Emerg Infect Dis. 2021;27(2):586–589.

7. Mohanan M, Malani A, Krishnan K, Acharya A. Prevalence of SARS-CoV-2 in Karnataka, India. JAMA. 2021;325(10):1001–1003. doi:10.1001/jama.2021.0332.

8. Govt of Telangana. MEDIA BULLETIN-COVID-19 Dated:05/10/2020 As of: 05/10/2020 (8PM). OFFICE OF THE DIRECTOR OF PUBLIC HEALTH AND FAMILY WELFARE, Hyderabad. [Accessed from https://covid19.telangana.gov.in/wp-content/uploads/2020/10/Media-Bulletin-05-10-2020.pdf on 05.10.2020 at 11:05 am]

9. Murhekar MV, Bhargava B, et al. SARS-CoV-2 seroprevalence among general population and healthcare workers in India, December 2020 - January 2021. Int J Infect Dis. 2021 May 19:S1201-9712(21)00442-2. doi: 10.1016/j.ijid.2021.05.040. Epub ahead of print. PMID: 34022338.

10. Murhekar MV, Bhargava B et al. Prevalence of SARS-CoV-2 infection in India: Findings from the national serosurvey, May-June 2020. Indian J Med Res. 2020 Jul & Aug;152(1 & 2):48–60. Doi: 10.4103/ijmr.IJMR_3290_20. PMID: 32952144; PMCID: PMC7853249.

11. https://www.the-scientist.com/features/sex-differences-in-immune-responses-to-viral-infection-68466

12. Population-based seroprevalence surveys of anti-SARS-CoV-2 antibody: An up-to-date review, IJID 2020

13. Leung JM, Yang CX, Tam A, et al. ACE-2 expression in the small airway epithelia of smokers and COPD patients: implications for COVID-19. Eur Respir J 2020; 55: 2000688 [https://doi.org/10.1183/13993003.00688-2020].

14. Kallathiyan, K., Velumani, A., Iyer, S., Sivapandi, K., & Velumani, A. (2020). COVID-19 seroprevalence study of an Indian Diagnostic Laboratory - Report on gender and age analysis. Asian Journal of Health Sciences, 6(2), Article ID 16. https://doi.org/10.15419/ajhs.v6i2.478

15. Anup Malani, Sabareesh Ramachandran, et.al. SARS-CoV-2 Seroprevalence in Tamil Nadu in October-November 2020. Preprint. Accessed from medRxiv 2021.02.03.21250949; doi: https://doi.org/10.1101/2021.02.03.21250949

16. Prajjval Pratap Singh, Rakesh Tamang, et.al. Estimation of real-infection and immunity against SARS-CoV-2 in Indian populations. Preprint. Accessed from medRxiv 2021.02.05.21251118; doi: https://doi.org/10.1101/2021.02.05.21251118

17. Prakash O, Solanki B, Sheth JK, et al. Assessing seropositivity for IgG antibodies against SARS-CoV-2 in Ahmedabad city of India: a cross-sectional study. BMJ Open 2021;11:e044101. doi: 10.1136/bmjopen-2020-044101

18. Khan SMS, Qurieshi MA, Haq I, et al. Seroprevalence of SARS-CoV-2 specific IgG antibodies in District Srinagar, northern India - A cross-sectional study. PLoS One. 2020;15(11):e0239303. Published 2020 Nov 11. doi:10.1371/journal.pone.0239303

